# Association of depression with anxiety, insomnia, social support, and social capital

**DOI:** 10.1101/2025.03.25.25324646

**Authors:** Feligno Alberto Barliza-Quintana, Carmen Cecilia Caballero-Domínguez, John Carlos Pedrozo-Pupo, Adalberto Campo-Arias

**Author notes:** **Correspondence:** Adalberto Campo-Arias, Universidad del Magdalena, Calle 29H3 No. 22-01, Santa Marta, Colombia (Postal code 470004). **Financing:** the Vice-Rector’s Office for Research of the University of Magdalena financed the project through resolution 0429 of 2022. **Ethics approval:** the Institutional Ethics Committee of the Universidad del Magdalena, Santa Marta, Colombia, approved the project according to minutes 005 of the virtual ordinary session of June 9, 2022. **Consent to participate:** Participants provided consent via online informed consent.

## Abstract

**Introduction:** The association of depression (depressive symptoms) with anxiety, insomnia, social support, and cognitive social capital has been studied in medical students. However, these associations have been analyzed independently, without adjusting or exploring possible interactions between them.

**Objective:** To evaluate the association of depression with anxiety, insomnia, social support, and cognitive social capital in medical students from Santa Marta, Colombia. Method: A cross-sectional study was designed, including medical students who reported symptoms of anxiety (GAD-7, Cronbach’s alpha of 0.91), insomnia (Athens Insomnia Scale, Cronbach’s alpha of 0.80), social support (Oslo-3, Cronbach’s alpha of 0.45), cognitive social capital (CSCS-5, Cronbach’s alpha of 0.77) and depression (PHQ-9, Cronbach’s alpha of 0.88). Using logistic regression, they measured the associations and interactions for depression.

**Results:** Three-hundred-nine students aged 18 and 39 participated (M=20.8, SD=2,7); 54.7% were women. 31.1% reported anxiety; 41.1%, insomnia; 57.3%, limited social support; 26.3%, poor cognitive social capital; and 39.5%, depression. Anxiety (OR=9.78, 95%CI 4.90-19.45), insomnia (OR=8.04, 95%CI 4.30-15.05), and the interaction between limited social support and poor cognitive social capital (OR=2.36, 95%CI 1.04-5.34) were significantly associated with depression (Hosmer-Lemeshow chi-square of 7.69, degree of freedom of 4, and p-value of 0.10, R-squared of Nagelkerke of 0.55).

**Conclusions:** Forty percent of medical students report symptoms of depression. Depression is significantly associated with anxiety, insomnia, social support, and cognitive social capital. New studies should explore other possible variables related to depression in Colombian medical students.

## INTRODUCTION

Depression is a common mental disorder that affects approximately more than 264 million people worldwide.^1^ Depression negatively affects the way people feel, think, and behave. Symptoms of a depressive episode include depressed mood, loss of interest or pleasure in activities, increased or decreased appetite, insomnia or hypersomnia, fatigue or loss of energy, feelings of worthlessness or guilt, difficulty concentrating, and thoughts of death.^2^

Depression is one of the most common health problems among university students, especially health sciences students.^3,4^ A similar situation is observed in Colombia, where studies reveal that the prevalence of depressive symptoms in university medical students significantly exceeds that observed in the general population, with frequencies between 25% and 52%.^5–7^

Early detection of depression in college students is crucial since most of the most common mental disorders manifest during this stage of life.^8^ Added to this is that the mental health of this population group has a significant impact on health services and mental health policies.^9^

This disorder deserves special attention because it affects students’ academic performance, dropout, and professional development.^10^ Unfortunately, depression in medical students also results in high rates of suicidal ideation, with reported prevalences of 11%.^11^ Furthermore, it may have implications for long-term patient care, as depressed medical students have been reported to be more cynical, less empathetic, and less willing to care for patients with chronic illnesses.^10^

Despite the alarming prevalence of depression in the university environment, medical students often refuse to consult, with care rates lower than 25%, with the diagnosis omitted and the appropriate treatment.^11^ Consequently, the need to implement strategies for students to seek professional help or attention in the university environment is imperative.^12^

It is essential to understand the complexities associated with depression. Frequent depression is associated with other mental health problems, such as anxiety or insomnia^13,14^ and, in addition, is influenced by social factors such as social support and cognitive social capital.^15^

In medical students, Alsheri et al.,^16^ out of 182 students evaluated depressive symptoms with the Patient Health Questionnaire (PHQ-9) and anxiety symptoms with the Generalized Anxiety Disorder Scale (GAD-7) and found that the presence of anxiety increased nine times the risk of depression, after adjusting for other variables. Likewise, Ibrahim et al.,^17^ in 450 students evaluated with the Hospital Anxiety and Depression Scale (HADS), found that the presence of anxiety increased three times the possibility of identifying a depressive episode.

However, Lepcha et al.,^18^ assessed depressive and anxious symptoms with the Hospital Anxiety and Depression Scale (HADS) and observed that the correlation between anxiety and depression scores was statistically low. Alzeghoul et al.,^19^ in 313 university students, applying the Beck Depression Inventory-II and the State-Trait Anxiety Questionnaire (STAI), found independence between the scores for anxiety and depression. The relationship between anxiety and depression is complex; Anxiety can predispose to depression, and vice versa, through interrelated biological and psychological mechanisms.^20,21^

Likewise, a strong association between insomnia and depression has been observed in medical students.^22^ Coico et al.,^23^ among 481 students, observed that students who reported insomnia were almost nine times more likely to have depression. Jeong et al.,^24^ in 120 students, reported that poor sleep quality increased the odds of depression more than eight times. However, Eller et al.,^25^ in 413 participants, found a significant relationship only between baseline insomnia and depressive symptoms. Insomnia can induce depression through a combination of neurobiological and stress reactivity factors.^26^ Hypersensitivity to stress makes it difficult to sleep and can precipitate depressive symptoms.^27–33^

Similarly, an inverse relationship has been found between perceived social support and depressive symptoms in medical students. Dhanoa et al.,^34^ among 177 students, found that participants who perceived a lack of support from their social networks, friends, and family were more than ten times more likely to experience depression than those who perceived high support. Likewise, Jeong et al.^24^ revealed that medical students with lower levels of perceived social support were ten times more likely to have depression. However, Yang et al.,^35^ in a sample of 457 students, observed independence between perceived social support and depression. Theoretically, social support is essential to face stressors and adversities. Therefore, limited social support may increase the risk of depression.^36^

Likewise, a significant correlation has been found between cognitive social capital and depression in university students; However, this association has been dismissed among medical students. Backhaus et al.,^37^ in 4,228 first-year university students from Europe, Asia, the Western Pacific, Latin America, and North America, reported that low levels of cognitive social capital increased the likelihood of depressive symptoms almost twofold. Similarly, Sotaquira et al.,^38^ in 216 university students, found that a low level of cognitive social capital approximately doubled the risk of clinically relevant depressive symptoms. However, Doĝan et al.,^39^ in 520 university students, found a lack of association between social information processing (an essential component of cognitive social capital)^40^ and depression. Low cognitive social capital may contribute to depression by impairing the sense of belonging and trust in social relationships.^41^ This situation can lead to feelings of isolation and hopelessness, which has been linked to the risk of depressive symptoms.^41,42^

This study, unlike previous research that has individually explored the relationship between depression and variables such as anxiety, insomnia, social support, and cognitive social capital in medical students,^14,16,17,24,43^ presents a holistic perspective of depression in this population by analyzing, adjusting, and exploring the possible interactions between these variables, which allows a realistic approach to the complex relationship between these factors.^44–46^ These findings may be helpful as more effective strategies to promote help-seeking in the presence of depressive symptoms.^46,47^

The objective of the study was to evaluate the association of depression with anxiety, insomnia, social support, and cognitive social capital in medical students from Santa Marta, Colombia.

## METHOD

### Participants

A cross-sectional analytical study was designed so all students of legal age, from the first to the tenth semester, were invited to participate. It was expected to have between 50 and 100 positive cases for the different variables of interest, given that this would allow adjustments to be made for up to ten variables at a rate of ten positive cases for each variable included in a logistic regression model.^48^

### Measurements

The research questionnaire investigated demographic information such as age, gender, stratum (income), origin, and semester. Additionally, students completed five measurement scales for anxiety, depression, social support, cognitive social capital, and depression.

Depression during the most recent two weeks was measured with the Patient Health Questionnaire (PHQ-9). Observable scores are between zero and 27; scores greater than nine were classified as high risk for depression (depressive symptoms with clinical significance).^49^ This instrument showed Cronbach’s alpha of 0.83 in Colombian medical students.^50^ In this group of students, the PHQ-9 achieved a Cronbach’s alpha of 0.88.

Anxiety was quantified with the Generalized Anxiety Disorder scale (GAD-7). The instrument includes seven items to evaluate symptoms during the last two weeks. Scores can be between zero and 21; scores greater than nine were classified as high risk for anxiety (anxiety symptoms with clinical significance).^51^ The GAD-7 showed Cronbach’s alpha of 0.93 in Colombian adults.^52^ In the present study, the GAD-7 presented Cronbach’s alpha of 0.91.

Insomnia was explored with the Athens Insomnia Scale (AIS). This scale consists of eight items divided into two dimensions: The first dimension groups five items to assess nighttime sleep during the last month and the second is made up of the remaining three to quantify daytime discomfort related to limited sleep. Scores can be between zero and 24. Scores above nine imply a high risk of insomnia. ^53^ In Colombian adults, the first dimension showed Cronbach’s alpha of 0.90 and the second 0.79.^54^ In the present investigation, the global Cronbach’s alpha was 0.80 (0.69 for the first dimension and 0.77 for the second).

Social support was quantified with the Oslo Social Support Scale-3 (OSSS-3). The instrument brings together three Likert-type items that explore the number of close people, people who show concern, and the ease of obtaining help from neighbors. The items can be used individually or as a construct (Cronbach’s alpha of 0.60). Scores can be between 3 and 14; the higher the score, the greater the social support. Scores less than ten were categorized as limited social support.^55^ In the present sample, the OSSS-3 presented Cronbach’s alpha of 0.45.

Cognitive social capital was measured with the Cognitive Social Capital Scale-5 (CSCS-5), derived from a seven-item version.^56^ This instrument consists of five items investigating current perceptions about the relationship with neighbors. The minimum score is zero, and the maximum is fifteen. Scores under five were categorized as low social capital (Cronbach’s alpha of 0.80 in Colombian adults).^57^ In the present study, the CSCS-5 showed Cronbach’s alpha of 0.77.

### Procedure

On the university campus, students completed an online version of the research questionnaire in a collective application in the classroom. Students of legal age who agreed to participate and signed the informed consent had a link to the Google form with all the items available. The questionnaire was presented item by item because the response grid format is less reliable and can induce errors.^58^ Completing the research questionnaire took an average of 20 minutes. The information was collected between September and November 2022.

### Data analysis

A descriptive analysis was carried out to establish the frequencies, percentages, means, and standard deviations according to the type of variables. Next, odds ratios (OR), with 95% confidence intervals (95%CI), were calculated to determine the association between the variables of interest and insomnia. Variables that showed probability values less than 0.20 and other recommendations presented by Greenland will be considered for adjustment and exploring interactions.^59^ For the acceptance of the model, the Hosmer-Lemeshow goodness of fit was calculated^,60^ and a probability value equal to or greater than 10% was expected.^48^ Finally, Nagelkerke’s R square was found to specify the percentage of variance of the variables included in the final model explained.^61^ The analysis was completed with the IBM-SPSS program.^62^

### Ethical aspects

The Institutional Ethics Committee of the Omitted University for Blind Evaluation approved this project according to minutes 005 of the virtual ordinary session of June 9, 2022. The students signed an informed consent.

## RESULTS

Three hundred nine students between 18 and 39 participated (M=20.8; SD=2.7). The most significant percentage of participants were between 18 and 20 years old, were female, came from an urban area, lived in low-income neighborhoods, were in a clinical semester, had limited social support, and reported sufficient cognitive social capital. See more demographic details in Table 1.

**Table 1.** Description of participating students (n=309).

| Variable | Frequency | % |
| --- | --- | --- |
| *Age (years)* |  |  |
| Between 18 and 20 | 166 | 53.7 |
| 21 or more | 143 | 46, 3 |
| *Gender* |  |  |
| Female | 169 | 54, 7 |
| Male | 140 | 45.3 |
| *Origin* |  |  |
| Urban | 266 | 86.9 |
| Rural | 43 | 13, 1 |
| *Residents in income neighborhoods* |  |  |
| Low | 194 | 62.8 |
| Middle or high | 115 | 37.2 |
| *Semester* |  |  |
| Primary (first to fifth) | 127 | 41.1 |
| Clinical (sixth to tenth) | 182 | 58.9 |
| *Social support* |  |  |
| Limited | 177 | 57, 3 |
| High | 132 | 42.7 |
| *Cognitive social capital* |  |  |
| Poor | 73 | 23.6 |
| Enough | 236 | 76, 4 |

On the other hand, 122 reported a high risk of depression (39.5%), 96 scored a high risk of anxiety (31.1%), 127 had a risk of insomnia (41.1%), 177 (57.3%) reported limited social support, and 77 (23.6%) reported poor cognitive social capital.

Raw associations were significant for gender, social support, cognitive social capital, anxiety, and insomnia and were considered to explore interactions and final adjustment. See raw OR and 95%CI values in Table 2. The most parsimonious, best-fitting model and highest value of Nagelkerke ‘s R square is presented in Table 3.

**Table 2.** Crude associations for sleepiness in Colombian medical students.

| Variable | OR | 95%CI |
| --- | --- | --- |
| Age over 20 years | 1.03 | 0.65-1.63 |
| Female gender | 1.87 | 1.17-2.99 |
| Urban origin | 1.60 | 0.80-3.21 |
| Residents in higher income neighborhood | 1.15 | 0.72-1.85 |
| Primary semester (first to fifth) | 1.11 | 0.70-1.76 |
| Limited social support | 1.77 | 1.10-2.83 |
| Low cognitive social capital | 3.07 | 1.79-5.28 |
| High risk of anxiety | 18.37 | 9.87-34.21 |
| High risk of insomnia | 14.43 | 8.26-25.23 |

**Table 3.** Adjusted association of depression with anxiety, social support, cognitive social capital, and insomnia.

| Variable | OR | 95%CI |
| --- | --- | --- |
| At high risk of anxiety | 9.76 | 4.90-19.45 |
| High risk of insomnia | 8.04 | 4.30-15.05 |
| Limited cognitive social capital*social support | 2.36 | 1.04-5.34 |

\*Interaction.
Hosmer-Lemeshow chi-square of 7.69; degree of freedom of 4, and p-value of 0.10. Nagelkerke R square of 0.55.

## DISCUSSION

In the present study, an association was observed between anxiety, insomnia, and depression. Furthermore, it was identified that the interaction between low social support and limited cognitive social capital was also related to the presence of depressive symptoms in university medical students in Santa Marta, Colombia.

In the current research, a strong relationship between anxiety and depression was identified. This finding is consistent with studies by Alsheri et al.,^16^ and Ibrahim et al.,^17^ which indicated that anxiety can significantly increase the risk of depressive symptoms in medical students. However, Lepcha et al.^18^ found independence between anxiety and depression.

The findings of the present study reveal a statistically significant association between insomnia and depression. This observation is similar to what has been reported in other investigations.^24–34^ For example, Coico et al.^23^ and Jeong et al.^24^ found that medical students with insomnia showed an increased risk of reporting depression. However, Eller et al.^25^ found independence between insomnia and depression.

The present research shows that limited cognitive social capital and poor social support doubled the probability of depression in medical students. These findings are consistent with Dhanoa et al.^34^ and Jeong et al.^24^ who documented a strong relationship between social capital and depression. Likewise, Backhaus et al.^37^ and Sotaquira et al.^38^ reported a statistically significant association between cognitive social capital and depression in college students. However, Yang et al.^35^ showed a lack of relationship between social support and depression. Doĝan et al.^39^ evidenced independence between social information processing (an essential component of cognitive social capital)^40^ and depression.

Discrepancies in findings may be attributed to critical differences in the demographic and cultural characteristics of the participating students and the instruments used in measuring the variables.^63^

These findings highlight the need to consider both emotional factors, such as anxiety and insomnia, and social factors, such as social support, and cognitive social capital, in understanding and addressing depression in medical students. It is crucial to implement screening programs for depression and other mental health problems in medical schools for early and timely detection.^13,64^ Additionally, teaching staff should be trained to identify signs and symptoms of depression and other mental health problems in students and suggest professional consultation for evaluation and treatment.^13,64^ Likewise, it is essential to create an environment in which students feel comfortable seeking help in the presence of depressive symptoms.^65^ Possibly, increasing mental health literacy can help identify predictors of depression in medical students and reduce the negative impact of depression in the academic context.^66^

One of the main strengths of this study is the exploration of the interaction between social support and cognitive social capital with depression, which highlights the depth of the analysis carried out and offers a complete vision of how these factors influence the mental health of medical students.^67^ However, this study has a significant limitation: The cross-sectional design limits establishing the causal directions between the variables analyzed. Additionally, by focusing exclusively on medical students from a specific location, the generalizability of the results to other populations or settings may be limited.^63^ Another relevant consideration is using a measurement scale with relatively low Cronbach’s alpha values (Oslo Social Support Scale-3), which could affect the reliability of the results in this measurement.^68^

## CONCLUSIONS

This study reveals that approximately 40% of medical students present symptoms of depression and highlights a significant association between depression and anxiety, insomnia, social support, and cognitive social capital. Future research should explore other variables potentially associated with depression in this specific population. Longitudinal studies will clarify the causal relationship when bidirectional relationships may exist. Likewise, using measurement scales with excellent reliability to quantify social support is advisable. Additionally, expanding the sample to include medical students from diverse geographic locations is recommended, which could improve the generalizability of the findings.

## Data Availability

All data produced in the present study are available upon reasonable request to the author.

## REFERENCES

1. World Health Organization. Depression: Fact sheet. Geneva: World Health Organization; 2020. 1. https://www.who.int/news-room/fact-sheets/detail/depression

2. American Psychiatric Association. Diagnostic and statistical manual of mental disorders: DSM-5-TR. 5th ed., text revision. Washington, DC: American Psychiatric Association; 2022.

3. Rotenstein LS, Ramos MA, Torre M, Segal JB, Peluso MJ, Guille C, et al. Prevalence of depression, depressive symptoms, and suicidal ideation among medical students: A systematic review and meta-analysis. JAMA. 2016;316(21):2214–36. 10.1001/jama.2016.17324

4. Nguyen M. Why medical school is depressing and what we should be doing about it. Aust Med Stud J. 2011;2(1):65–8.

5. Quintero MA, García CC, Jiménez VLG, Ortiz TML. [Characterization of depression in young university students]. Univ Psychol. 2004;3(1):17–26.

6. Guavita P, Sanabria P. [D epressive symptoms prevalence, in one medical student population. Universidad Militar Nueva Granada School of Medicine. Bogota-Colombia]. Rev Fac Med. 2006;54:76–87.

7. Miranda CA, Gutiérrez JC, Bernal F, Escobar CA. [Prevalence of depression medical students of the Universidad del Valle]. Rev Colomb Psiquiatr. 2000;29(3):251–60.

8. Kessler RC, Bromet EJ. The epidemiology of depression across cultures. Annu Rev Public Health. 2013;34:119–38. 10.1146/annurev-publhealth-031912-114409

9. Viñas F, Villar E, Caparros B, Juan J, Cornella M, Perez I. Feelings of hopelessness in a Spanish university population: Descriptive analysis and its relationship to adapting to university, depressive symptomatology and suicidal ideation. Soc Psychiatry Psychiatr Epidemiol. 2004;39:326–34. 10.1007/s00127-004-0756-2

10. Puthran R, Zhang MW, Tam WW, Ho RC. Prevalence of depression amongst medical students: A meta-analysis. Med Educ. 2016;50(4):456–68. 10.1111/medu.12962

11. Dyrbye LN, Thomas MR, Massie FS, Power DV, Eacker A, Harper W, et al. Burnout and suicidal ideation among US medical students. Ann Intern Med. 2008;149(5):334–41. 10.7326/0003-4819-149-5-200809020-00008

12. Tjia J, Givens JL, Shea JA. Factors associated with undertreatment of medical student depression. J Am Coll Health. 2005;53(5):219–24. 10.3200/JACH.53.5.219-224

13. Mirza AA, Baig M, Beyari GM, Halawani MA, Mirza AA. Depression and anxiety among medical students: A brief overview. Adv Med Educ Pract. 2021;12:393–8. 10.2147/amep.s302897

14. Barakat D, Elwasify M, Elwasify M, Radwan D. Relation between insomnia and stress, anxiety, and depression among Egyptian medical students. Middle East Curr Psychiatry. 2016;23(3):119–27.

15. Lee SH, Lee H, Yu S. Effectiveness of social support for community-dwelling elderly with depression: A systematic review and meta-analysis. Healthcare. 2022;10(9):1598. 10.3390/healthcare10091598

16. Alshehri A, Alshehri B, Alghadir O, Basamh A, Alzeer M, Alshehri M, et al. The prevalence of depressive and anxiety symptoms among first-year and fifth-year medical students during the COVID-19 pandemic: A cross-sectional study. BMC Med Educ. 2023;23(1):411. 10.1186/s12909-023-04387-x

17. Ibrahim N, Dania AK, Lamis EK, Ahd AH, Asali D. Prevalence and predictors of anxiety and depression among female medical students in King Abdulaziz University, Jeddah, Saudi Arabia. Iran J Public Health. 2013;42(7):726.

18. Lepcha C, Kumar S, Mujeeb N, Sharma S. Anxiety and depression among medical undergraduate students and their sociodemographic correlates in Sikkim: A cross-sectional study. Natl J Physiol Pharm Pharmacol. 2022;12(6):899–902. 10.5455/njppp.2022.12.11420202105122021

19. Alzeghoul EA, Masten WG, Toy Caldwell-Colbert A, Williams V, Gadzella B, Helton JR, Ball SE. Empirical evidence for discriminating anxiety from depression in college students. Iran J Psychol. 2001;22(1):73–78. 10.1080/03033910.2001.10558265

20. Baldwin DS, Evans DL, Hirschfeld RM, Kasper S. Can we distinguish anxiety from depression? Psychopharmacol Bull. 2002;36:158–65.

21. Stein DJ, Westenberg HG, Liebowitz MR. Social anxiety disorder and generalized anxiety disorder: Serotonergic and dopaminergic neurocircuitry. J Clin Psychiatry. 2002;63:12–9.

22. Chang PP, Ford DE, Mead LA, Cooper-Patrick L, Klag MJ. Insomnia in young men and subsequent depression: The Johns Hopkins Precursors Study. Am J Epidemiol. 1997;146(2):105–14. 10.1093/oxfordjournals.aje.a009241

23. Coico-Lama AH, Diaz-Chingay LL, Castro-Diaz SD, Céspedes-Ramírez ST, Segura-Chavez LF, Soriano-Moreno AN. [Association between sleep disturbances and mental health problems in medical students during the COVID-19 pandemic]. Educ Med. 2022;23(3):100744. 10.1016/j.edumed.2022.100744

24. Jeong YJ, Kim JY, Ryu JS, Lee KE, Ha EH, Park HS. The associations between social support, health-related behaviors, socioeconomic status and depression in medical students. Epidemiol Health. 2010;32:1–8. 10.4178%2Fepih%2Fe2010009

25. Eller T, Aluoja A, Vasar V, Veldi M. Symptoms of anxiety and depression in Estonian medical students with sleep problems. Depress Anxiety. 2006;23(4):250–6. 10.1002/da.20166

26. Vargas I, Perlis ML. Insomnia and depression: Clinical associations and possible mechanistic links. Curr Opin Psychol. 2020;34:95–9. 10.1016/j.copsyc.2019.11.004

27. Haeffel GJ, Grigorenko EL. Cognitive vulnerability to depression: Exploring risk and resilience. Child Adolesc Psychiatr Clin North Am. 2007;16(2):435–48. 10.1016/j.chc.2006.11.005

28. Morin CM, Rodrigue S, Ivers H. Role of stress, arousal, and coping skills in primary insomnia. Psychosom Med. 2003;65(2):259–67. 10.1097/01.psy.0000030391.09558.a3

29. Pigeon WR, Perlis ML. Insomnia and depression: Birds of a feather? Int J Sleep Disord Res. 2007;1(3):84–93.

30. Adrien J. Neurobiological bases for the relation between sleep and depression. Sleep Med Rev. 2002;6(5):341–51. 10.1053/smrv.2001.0200

31. Finan PH, Smith MT. The comorbidity of insomnia, chronic pain, and depression: Dopamine as a putative mechanism. Sleep Med Rev. 2013;17(3):173–83. 10.1016/j.smrv.2012.03.003

32. Palagini L, Biber K, Riemann D. The genetics of insomnia–evidence for epigenetic mechanisms? Sleep Med Rev. 2014;18(3):225–35. 10.1016/j.smrv.2013.05.002

33. Levenson JC, Kay DB, Buysse DJ. The pathophysiology of insomnia. Chest. 2015;147(4):1179–92. 10.1378/chest.14-1617

34. Dhanoa S, Oluwasina F, Shalaby R, Kim E, Agyapong B, Hrabok M, et al. Prevalence and correlates of likely major depressive disorder among medical students in Alberta, Canada. Int J Environ Res Public Health. 2022;19(18):11496. 10.3390/ijerph191811496

35. Yang D, Oral E, Kim J, Craft T, Moore MB. Depression and perceived social support in Asian American medical students. J Racial Ethn Health Disparities. 2021;1–11. 10.1007/s40615-021-01043-2

36. Cohen SR, Wills TA. Stress, social support, and the buffering hypothesis. Psychol Bowl. 1985;98(2):310–57.

37. Backhaus I, Varela AR, Khoo S, Siefken K, Crozier A, Begotaraj E, et al. Associations between social capital and depressive symptoms among college students in 12 countries: results of a cross-national study. Front Psychol. 2020;11:644. 10.3389/fpsyg.2020.00644

38. Sotaquirá L, Backhaus I, Sotaquirá P, Pinilla-Roncancio M, González-Uribe C, Bernal R, et al. Social capital and lifestyle impacts on mental health in university students in Colombia: An observational study. Front Public Health. 2022;10:840292. 10.3389/fpubh.2022.840292

39. Doĝan T, Çetin B. The investigation of relationship between social intelligence, depression and some variables at university students. J Human Sci. 2008;5:2.

40. Nahapiet J, Ghoshal S. Social capital, intellectual capital, and the organizational advantage. Acad Manag Rev. 1998;23(2):242–66. 10.5465/amr.1998.533225

41. Bassett E, Moore S. Social capital and depressive symptoms: the association of psychosocial and network dimensions of social capital with depressive symptoms in Montreal, Canada. Soc Sci Med. 2013;86:96–102. 10.1016/j.socscimed.2013.03.005

42. Cohen-Cline H, Beresford SA, Barrington W, Matsueda R, Wakefield J, Duncan GE. Associations between social capital and depression: A study of adult twins. Health Place. 2018;50:162–7. 10.1016/j.healthplace.2018.02.002

43. Mayer FB, Santos IS, Silveira PS, Lopes MHI, de Souza ARND, Campos EP, et al. Factors associated to depression and anxiety in medical students: A multicenter study. BMC Med Educ. 2016;16(1):282. 10.1186/s12909-016-0791-1

44. Vargas SM. [Factors that influence depression in university students: Systematic review]. Conrado. 2021;17(82):387–94.

45. Dafaalla M, Farah A, Bashir S, Khalil A, Abdulhamid R, Mokhtar M, et al. Depression, anxiety, and stress in Sudanese medical students: A cross-sectional study on role of quality of life and social support. Am J Educ Res. 2016;4(13):937–42.

46. Moir F, Yielder J, Sanson J, Chen Y. Depression in medical students: Current insights. Adv Med Educ Pract. 2018;9:323–33. 10.2147%2FAMEP.S137384

47. Thompson D, Goebert D, Takeshita J. A program for reducing depressive symptoms and suicidal ideation in medical students. Acad Med. 2010;85(10):1635–9. 10.1097/acm.0b013e3181f0b49c

48. Hosmer DW, Lemeshow S, Sturdivant RX. Applied logistic regression. New York: John Wiley & Sons; 2013.

49. Kroenke K, Spitzer RL, Williams JB. The PHQ-9: Validity of a brief depression severity measure. J Gen Intern Med. 2001;16(9):606–13. 10.1046/j.1525-1497.2001.016009606.x

50. Cassiani-Miranda CA, Vargas-Hernández MC, Pérez-Anibal E, Herazo-Bustos MI, Hernández-Carrillo M. [Reliability and dimensionality of PHQ-9 in screening symptoms of depression among health science students in Cartagena, 2014]. Biomedica. 2016;37(1):112–20. 10.7705/biomedica.v37i0.3221

51. Spitzer RL, Kroenke K, Williams JB, Löwe B. A brief measure for assessing generalized anxiety disorder: The GAD-7. Arch Intern Med. 2006;166(10):1092–7. 10.1001/archinte.166.10.1092

52. Monterrosa-Blanco A, Cassiani-Miranda CA, Scoppetta O, Monterrosa-Castro Á. Generalized anxiety disorder scale (GAD-7) has adequate psychometric properties in Colombian general practitioners during COVID-19 pandemic. Gen Hosp Psychiatry. 2021;70:147–8. 10.1016/j.genhosppsych.2021.03.013

53. Soldatos CR, Dikeos DG, Paparrigopoulos TJ. Athens Insomnia Scale: Validation of an instrument based on ICD-10 criteria. J Psychosom Res. 2000;48(6):555–60. 10.1016/s0022-3999(00)00095-7

54. Campo-Arias A, Caballero-Domínguez CC, Pedrozo-Pupo JC. [Reliability and validity of the Athens Insomnia Scale among Colombian People Recovered from COVID-19]. Rev Colomb Neumol. 2024;36(2):18–24. 10.30789/rcneumologia.v36.n2.2024.624

55. Dalgard OS, Dowrick C, Lehtinen V, Vazquez-Barquero JL, Casey P, Wilkinson G, et al. Negative life events, social support and gender difference in depression. A multinational community survey with data from the ODIN study. Soc Psychiatry Psychiatr Epidemiol. 2006;41:444–51. 10.1007/s00127-006-0051-5

56. Martin KS, Rogers BL, Cook JT, Joseph HM. Social capital is associated with decreased risk of hunger. Soc Sci Med. 2004;58(12):2645–54. 10.1016/j.socscimed.2003.09.026

57. Campo-Arias A, Caballero-Domínguez CC, Pedrozo-Pupo JC. Internal consistency and dimensionality assessment of the Cognitive Social Capital Scale among Colombian adults. Med Clin Soc. 2024;8(1):75–83. 10.52379/mcs.v8i1.363

58. Revilla M, Toninelli D, Ochoa C. An experiment comparing grids and item-by-item formats in web surveys completed through PCs and smartphones. Telemat Informat. 2017;34(1):30–42. 10.1016/j.tele.2016.04.002

59. Greenland S. Modeling and variable selection in epidemiologic analysis. Am J Public Health. 1989;79(3):340–9. 10.2105/AJPH.79.3.340

60. Hosmer DW, Lemeshow S. Goodness of fit tests for the multiple logistic regression model. Commun Stat. 1980;9(10):1043–69. 10.1080/03610928008827941

61. Nagelkerke NJ. A note on a general definition of the coefficient of determination. Biometrika. 1991;78(3):691–2.

62. IBM Corp. IBM SPSS Statistics for Windows, Version 24.0. Armonk, NY: IBM Corp. 2016.

63. Wang X, Cheng Z. Cross-sectional studies: Strengths, weaknesses, and recommendations. Chest. 2020;158(1):S65–71. 10.1016/j.chest.2020.03.012

64. Leiva-Nina M, Indacochea-Cáceda S, Cano LA, Medina Chinchon M. [Association between anxiety and depression in medical students at the Ricardo Palma University during the year 2021]. Rev Fac Med Hum. 2022;22(4):735–742. https://revistas.urp.edu.pe/index.php/RFMH/article/view/4842

65. Almanasef M. Mental health literacy and help-seeking behaviours among undergraduate pharmacy students in Abha, Saudi Arabia. Risk Manag Health Policy. 2021;1281–1286.

66. Zhong Y, Schroeder E, Gao Y, Guo X, Gu Y. Social support, health literacy and depressive symptoms among medical students: an analysis of mediating effects. Int J Environ Res Public Health. 2021;18(2):633.

67. Talaei A, Ardani AR, Saghebi A. A survey of depression among Iranian medical students and its correlation with social support and satisfaction. J Pakistan Psychiatr Soc. 2008;5(2):90–9.

68. Kalkbrenner MT. Alpha, omega, and H internal consistency reliability estimates: Reviewing these options and when to use them. Counsel Outcome Res Eval. 2023;14(1):77–88. 10.1080/21501378.2021.1940118

